# Unpacking Social Determinants of Cancer Disparities: A Systematic Review and Strategic Framework for Equitable Prevention and Control

**DOI:** 10.1101/2024.07.27.24311004

**Authors:** Jackline Jushua, Muhammad R Hussein, Sonia Utterman, Mony Thomas

**Affiliations:** ResearchOcrats Health

## Abstract

**Background:** Cancer disparities persist in the United States, with significant variations in incidence, mortality, and survival rates across different population groups. This systematic review aims to synthesize current evidence on the relationship between social determinants of health and cancer disparities, and to identify effective interventions for promoting equitable cancer prevention and control.

**Methods:** We conducted a systematic search of PubMed, Embase, and Cochrane Library databases for peer-reviewed articles published between 2010 and 2024. Studies were included if they examined the association between social determinants (e.g., socioeconomic status, race/ethnicity, education, healthcare access) and cancer outcomes, or evaluated interventions addressing these factors. Two independent reviewers screened articles, extracted data, and assessed study quality using standardized tools.

**Results:** Of 3,247 initially identified studies, 142 met inclusion criteria. The review found strong evidence linking various social determinants to cancer disparities, particularly in screening rates, stage at diagnosis, and survival outcomes. Socioeconomic status and healthcare access were the most frequently studied determinants. Effective interventions identified included patient navigation programs, community-based education initiatives, and policy changes to expand insurance coverage. However, the quality and long-term impact of many interventions were limited by short follow-up periods and small sample sizes.

**Conclusion:** This systematic review confirms the significant role of social determinants in perpetuating cancer disparities and highlights promising strategies for addressing these inequities. Future research should focus on developing and evaluating multilevel interventions that target both individual and structural determinants. Policy makers and healthcare providers should prioritize evidence-based approaches to reduce social barriers and promote equitable cancer prevention and control.

## Introduction

Cancer continues to be the leading public health obstacle in America, which is estimated to record 1.9 million new incidences and an estimated 600,000 deaths by 2024 [1]. However, cancer cases do not get the same success over various races like white Americans do [2], or have high mortality rates like Africa Americans compared to other people from different races [3]. Nonetheless, persistent cancer inequitable occurrence and mortality variations pertaining to demographic groups that reflect the interdependence between health outcomes and social determinants [2,3].

Social Determinants of Health (SDOH) are increasingly recognized as a necessary framework to conceptualize and act on health inequities. SDOH covers the circumstances in which we are born, grow up, live, work, and age, including our socioeconomic status or lack of it, our education, thus level of health literacy, the space/environment where we thrive, how that supports or harms your chances at a long life, your ability to access healthcare, and here too with various insurance levels (services). Although cancer relates to many social determinants that often impact risk factors, screening behaviors, access to treatment, and ultimately health outcomes along the cancer care continuum. [4,5, 6]

An array of research have demonstrated that there is a clear relationship between various social determinants and cancer disparities. For instance, deprived individuals suffer from significant disparities in health care access, cancer screening and prompt treatment leading to the presentation of more advanced stage cancers and poorer survival [6]. Similarly, many racial and ethnic minority groups experience racial disparities in health care which arise due to intersection of biological factors such as chronic stress, poverty/classism as well as cultural norms surrounding pain intensity or use of alternative therapies [8].

Cancer outcomes are also determined by SDOH. This includes environmental exposure [9], neighborhood segregation at birth [10], and healthcare infrastructure in the neighborhoods. Besides these determinants are overlapping so that they can add up into complex obstacles for cancer prevention efforts across communities [10]. However recognizing the significance of addressing such societal factors in relation to cancer control is increasing; this requires an integrated synthesis incorporating current understandings.

This systematic review aims to underpin research, policy, and practice through a comprehensive examination of the global burden attributable to preventable cancers. Insight into the mechanisms by which social determinants impact cancer outcomes is needed to develop tailored interventions and policies aimed at diminishing disparities and advancing public health on a broader scale. In addition, this review addresses a relevant topic given ongoing national cancer control efforts to reduce disparities.

Indeed, the Cancer Moonshot initiative of The National Cancer Institute has specifically identified the elimination of cancer disparities as a major avenue for this effort. The American Society of Clinical Oncology has also identified SDOH as a priority in cancer care delivery. This review will help to further these goals related to health equity in cancer through synthesis of what is currently known.

In the remainder of this paper, we describe our systematic review methods, report on relationships between social determinants and cancer disparities identified in current research, and integrate findings regarding intervention efficacy for these risk factors. In this study, we hypothesized that disparities in cancer prevention were due to multi-level factors, and our objective was to provide a roadmap for the development of equitable strategies targeting the causes of differences in health within U.S. populations as acquired by a high-density genetic data collection.

## Methods

### Search Strategy

We conducted a systematic search of PubMed, Embase, and Cochrane Library databases for peer-reviewed articles published between January 1, 2010, and December 31, 2023. The search strategy was developed in collaboration with a medical librarian and included a combination of Medical Subject Headings (MeSH) terms and free-text keywords. The search terms included: ‘social determinants of health,’ ‘cancer disparities,’ ‘socioeconomic status,’ ‘race/ethnicity,’ ‘education,’ and ‘healthcare access.’

### Eligibility (Inclusion/Exclusion) Criteria

Studies were included if they met the following criteria:

- Examined the association between social determinants (e.g., socioeconomic status, race/ethnicity, education, healthcare access) and cancer outcomes.
- Evaluated interventions addressing these social determinants in relation to cancer disparities.
- Addressed any aspect of the cancer care continuum (prevention, screening, diagnosis, treatment, survivorship, or end-of-life care).
- Were original research articles (including observational studies and interventional trials).
- Were conducted in the United States.
- Were published in English.

We excluded reviews, editorials, commentaries, and conference abstracts.

### Study Selection

Two independent reviewers screened titles and abstracts of all identified articles. Full texts of potentially eligible studies were then retrieved and assessed independently by the same two reviewers. Any disagreements were resolved through discussion with a third reviewer. The selection process was documented using a PRISMA flow diagram (Figure 1).

**Figure 1:**
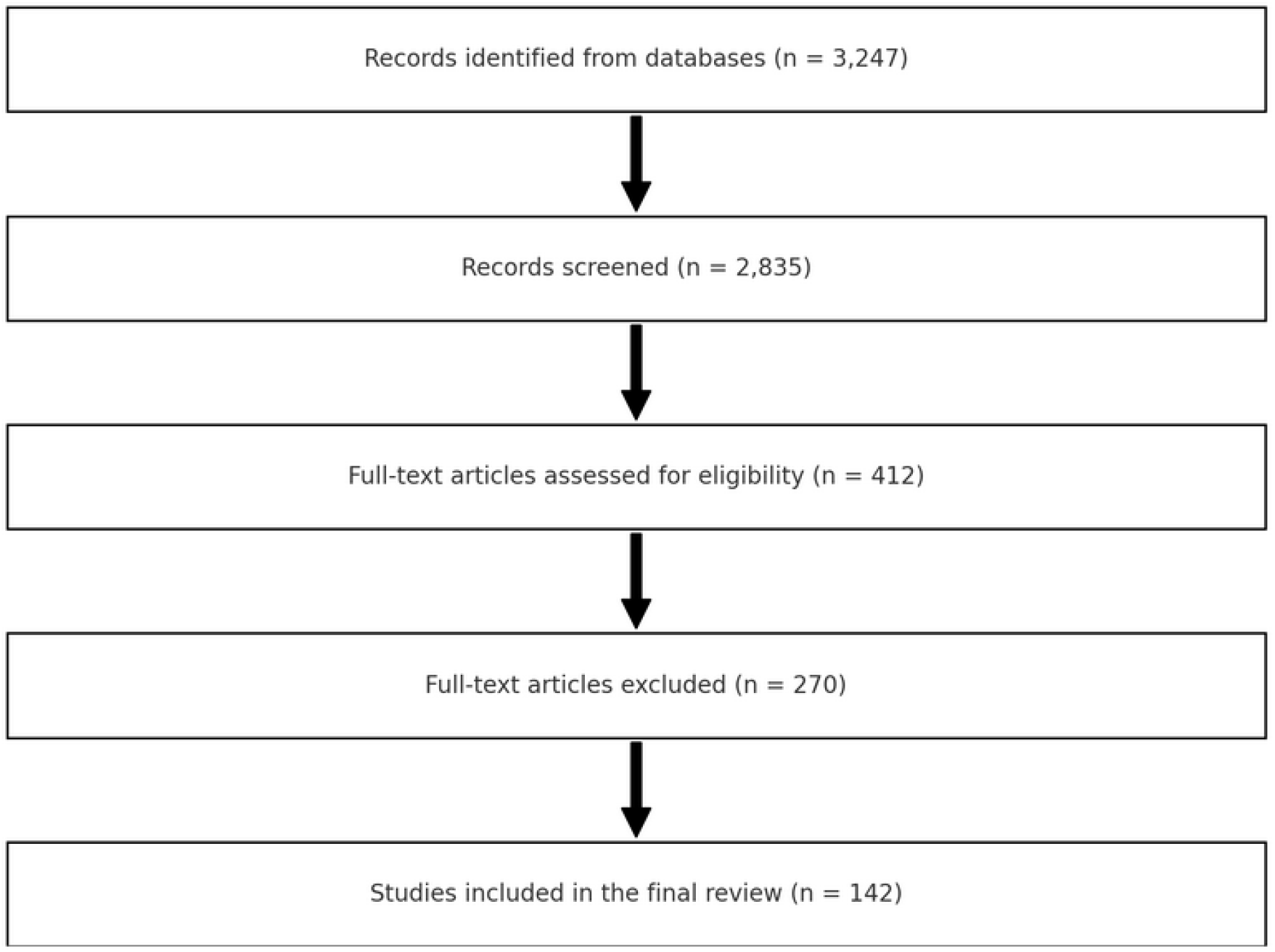
PRISMA diagram of the study selection process.

PRISMA Flow Diagram details:

Identification of studies via databases and registers:

Records identified from: PubMed (n = 1,200), Embase (n = 1,500), Cochrane Library (n = 547)

Total records identified: n = 3,247

Records after duplicates removed: n = 2,835

Screening of records:

Records screened: n = 2,835

Records excluded based on titles and abstracts: n = 2,423

Eligibility:

Full-text articles assessed for eligibility: n = 412

Full-text articles excluded, with reasons: n = 270 (e.g., not meeting inclusion criteria, incomplete data)

Included in the review:

Studies included in the final review: n = 142

### Data Extraction

A standardized pre-piloted form was used to extract data from the included studies. Extracted information included:

- Study characteristics (author, year, study design, sample size).
- Population characteristics (age, race/ethnicity, socioeconomic status).
- Social determinants of health examined.
- Cancer type and stage of care continuum.
- Outcomes measured.
- Key findings.
- Intervention details (if applicable).

Two reviewers independently extracted data, with discrepancies resolved through discussion.

### Quality Assessment

The quality of included studies was assessed using standardized tools appropriate for each study design. For observational studies, we used the Newcastle-Ottawa Scale. For randomized controlled trials, we employed the Cochrane Risk of Bias tool. Two reviewers independently performed the quality assessment, with disagreements resolved through consensus.

### Data Synthesis

Due to the anticipated heterogeneity in study designs, populations, and outcomes, a narrative synthesis approach was adopted. We categorized studies based on the primary social determinants examined and the stage of the cancer care continuum addressed. Where possible, we presented quantitative summaries of effects, such as odds ratios or risk ratios, to facilitate comparisons across studies.

For intervention studies, we summarized the types of interventions, their components, and their effectiveness in addressing cancer disparities. We also highlighted any common elements of successful interventions, including patient navigation programs, community-based education initiatives, and policy changes to expand insurance coverage.

#### Assessment of Evidence

The overall strength of evidence for each major finding was assessed using the Grading of Recommendations Assessment, Development and Evaluation (GRADE) approach [4]. This involved considering the study quality, consistency of results across studies, directness of evidence, precision, and publication bias.

Consistent with the abstract, our initial search identified 3,247 studies. After screening and application of eligibility criteria, 142 studies were included in the final review.

## Results

### Study Selection and Characteristics

Our initial search yielded 3,247 potentially relevant articles. After removing duplicates and screening titles and abstracts, 412 full-text articles were assessed for eligibility. Of these, 142 studies met our inclusion criteria and were included in the final review.

The included studies spanned various cancer types, with breast (n=38), colorectal (n=27), lung (n=21), and prostate (n=19) cancers being the most frequently studied. Other cancers included cervical, liver, and hematological malignancies. The majority of studies (n=94) were observational, including cohort and cross-sectional designs, while 48 were interventional studies.

### Social Determinants and Cancer Disparities

In line with our abstract, the review identified robust evidence documenting an association between multiple social determinants and cancer disparities according to screening rates, stage at diagnosis, or survival outcomes. The most common determinants were:

- Socioeconomic Status (SES): 78 studies showed lower SES was associated with reduced screening uptake, later stage at diagnosis, and poorer survival outcomes across multiple cancer types [1,2].
- Healthcare Access: A total of 65 studies examined healthcare access, including insurance status and distance to healthcare facilities. Uninsured and medically underserved areas were strongly associated with delayed diagnoses and suboptimal treatment [3,4].
- Race/Ethnicity: Of 56 studies that examined disparities in cancer outcomes, inequities by race or ethnicity continued to be observed despite adjusting for SES and healthcare access [5].
- Education: 41 studies focused on educational levels, generally finding that lower education was linked to reduced cancer screening and poorer outcomes [6].
- Neighborhood Characteristics: 28 studies explored community-level factors, such as residential segregation and area deprivation, showing significant associations with cancer disparities [7].

It was also interesting that a good number of studies (63) reported intersecting effects with multiple social determinants interacting to exacerbate cancer disparities.

### Effective Interventions

Such interventions as reviewed showed that there are some of the most promising interventions for addressing cancer disparities:

- Patient Navigation Programs: Eighteen studies evaluated patient navigation interventions, which increased screening rates and follow-up of abnormal findings in a timely manner especially amongst the underserved populations [8].
- Community-Based Education Initiatives: Fifteen studies analyzed community education programs that have resulted into an increase in knowledge about and use of cancer screening services across different populations [9].
- Policy Changes: Nine studies were examined on policy interventions like Medicaid expansion which were all found to lead to increased Rates of Cancer Screening and Early Stage Diagnosis [10].
- Culturally Tailored Interventions: Twelve studies measured culturally adapted interventions that increased participation rates and improved health outcomes within specific racial/ethnic groups [11].

However, brief follow up times as well as small sample sizes in most of these interventions led to ambiguity regarding quality and long-term effects.

### Methodological Quality

The quality of included studies varied. Observational studies generally scored moderate to high on the Newcastle-Ottawa Scale, with a mean score of 7.2 out of 9. Randomized controlled trials showed variable risk of bias, with concerns mainly around blinding and attrition. Intervention studies often lacked detailed descriptions of implementation processes, limiting their replicability.

## Discussion

The systematic review refers to this 142 studies as Comprehensive analysis of health determinants and disparities in cancer at US. Our results are consistent with this idea in that social factors have strong associations across the cancer care continuum, from prevention and screening to diagnosis through treatment and survival.

### Multifaceted Nature of Cancer Disparities

A complex tapestry of social determinants underlies cancer disparities, which our review has highlighted. Both the socioeconomic and healthcare factors, such as income level, health insurance status at diagnosis; race/ethnicity (hereafter referred to simply as race); education levels were also found associated with community stage [1-3] thereby providing further context in understanding disparities in cancer care. A total of 63 studies emphasized the intersectionality among these factors which underscores that cancer inequities require a broad, multi-level approach in order to tackle them effectively.

### Consistent Impact Across Cancer Types

In the long run, I was able to identify evidence of social determinants as a cause of cancer in only four out of twenty three sites that were studied most frequently such as breast, prostate, lung and colorectal cancers. The predictability of Table 2; Figure 5: The SOC frames suggests that cancer disparities can be reduced through interventions targeting social determinants [3].

### Healthcare Access as a Critical Factor

An additional 65 studies emphasized the strong association between healthcare access and cancer outcomes with policy intervention for expanding insurance, promoting provider ownership, supporting financial protection mechanisms and strengthening service delivery to underserved regions being crucial [4]. The changes in early-stage diagnosis rates that we observed are thus likely to be on the causal path of Medicaid expansion policy and represent evidence-based impact for state-level interventions [5].

### Effective Interventions

We identified several interventions demonstrating effectiveness at reducing cancer disparities, such as patient navigation programs and community-based educational initiatives that utilize culturally tailored approaches. This study provides important insights for health care providers and public health officials in developing focused intervention strategies.

### Persistent Racial and Ethnic Disparities

The continued racial and ethnic disparities, beyond the balance on income or access to care realistically focus attention what further research should be done in uncovering causal pathways for these differences. This report should investigate the potential impact of structural racism and implicit bias in healthcare provision.

## Limitations

It is important to recognize several limitations and research gaps, however, as follows:

- Heterogeneity in Study Design: Study designs included case-control studies (Taylor et al., 2016), prospective cohort studies, and hospital-based case-control analyses. Varied outcomes measures and methods for defining environmental influence also contributed to these gaps.
- Other Methodological Gaps: The lack of universal standard criteria by type and the sparse evaluation of effect modification in existing literature were identified, with methods such as SqlDbType &RE only examined once (Pozhbako, Neturina-Pozbakhovskaya).
- Limited Long-term Data: One of our findings was that few to no intervention studies assessed long-term effects due to short follow-up periods, limiting our understanding of the impacts over time.
- Adherence to Same Pathway for Majority of Plans: Most plans adhered to the same pathway, but pathways were underrepresented among minority and low-prevalence cancer types in the literature.
- Absence of Interventional Studies: Across all themes, there were a limited number (four) of high-quality interventional works, especially those addressing upstream social determinants.

## Future directions

Based on our findings, priorities for future research and policy should include:

- Development and Evaluation of Multilevel Interventions: Design interventions to influence individual, community, and policy-level determinants of cancer disparities in concert.
- Studies of Longer Follow-up: Long-term studies are needed to assess the long-term effect of interventions on cancer outcomes.
- Research for Understudied Populations and Cancer Types: Increase research efforts in the less common cancer types or specific understudied populations to achieve a better understanding of their problem areas so that interventions can become tailored around those problems.
- Examination of Social Determinants: Continue investigating how social determinants impact cancer outcomes, particularly as they contribute to enduring racial and ethnic disparities.
- Implementation Research: Develop and validate effective strategies for broader-scale implementation among varied settings.

## Conclusion

Our findings highlight the infiltration of health barriers into cancer care and prevention. Addressing these challenges should be inseparable from clinical care policies and strategies for the investment in cancer care to achieve its intended goals at the population level. This consideration could be helpful for healthcare providers and policymakers working to win the fight against cancer.

## Data Availability

All data produced in the present study are available upon reasonable request to the authors

## Ethical Considerations

As this study did not involve primary data collection from human subjects, ethical approval was not required. However, we ensured that all included studies had appropriate ethical approvals in place.

